# Antibody profiling reveals gender differences in response to SARS-COVID-2 infection

**DOI:** 10.1101/2021.06.01.21258175

**Authors:** Lia Tsverava, Nazibrola Chitadze, Gvantsa Chanturia, Merab Kekelidze, David Dzneladze, Paata Imnadze, Amiran Gamkrelidze, Vincenzo Lagani, Zaza Khuchua, Revaz Solomonia

**Author notes:** To whom correspondence should be addressed (E.mail).

## Abstract

The recent emergence of the novel severe acute respiratory syndrome coronavirus 2 (SARS-CoV-2) has led to an ongoing global COVID-19 pandemic and public health crisis. Detailed study of human immune response to SARS-COVIS-2 infection is the important topic for a successful treatment of this disease. Our study was aimed to characterize immune response on the level of antibody profiling in convalescent plasma of patients in Georgia. Antibodies against the following SARS-COV-2 proteins were studied: nucleocapsid and various regions of Spike (S) protein: S1, S2 and Receptor binding domain (RBD). Convalescent plasma of patients 6-8 weeks after initial confirmation of SARS-COV-2 infection were tested. Nearly 80% out of 154 patients studied showed presence of antibodies against nucleocapsid protein. The antibody response to three fragments of S protein was significantly less and varied in the range of 20-30%. Significantly more females as compared to males were producing antibodies against S1 fragment, whereas the difference between genders by the antibodies against nucleocapsid protein and RBD was statistically significant only by one-tailed Fisher exact test. There were no differences between the males and females by antibodies against S2 fragment. Thus, immune response against some viral antigens are stronger in females and we suggest that it could be one of the factors of less female fatality after SARS-COVID-2 infection.

## Introduction

The emergence of SARS-CoV-2 resulted in over 170 million (as of May 30,2021)infections and more than 3.5 million deaths worldwide. A growing body of evidence suggests sex differences in the clinical outcomes of coronavirus disease. Large-scale data analysis of global data suggests that males face higher odds of both intensive therapy unit (ITU) admission and death from COVID-19 compared to females [1-3]. The situation in Georgia is similar to global statistics. According to the available official statistics in Georgia for 15^th^ of April 2021 (see https://stopcov.ge/en) the number of SARS-COV-2 infected females were more than males (57% vs 43%) but the lethality was significantly higher in mans (56%vs44%, Chi2 test P<2.2e-16).

To elucidate the immune responses against SARS-CoV-2 infection in men and women, we performed antibody profiling of convalescent plasma from patients in Georgian Republic. In particular, immunoblot analysis was performed to identify possible associations between gender and the presence of proteins in COVID-19 patients’ blood plasma. We have studied the presence of antibodies against the various fragments of Spike (S) protein and nucleocapsid protein (NCP). This later one is an internal viral protein and not exposed on the surface of virion particles [4,5]. Antibodies against NCP thus lack neutralizing capacities. However, in patients with severe acute respiratory syndrome (SARS) the antibodies were mainly against NCP [6] and it is suggested that their production might reflect the strength of T-helper cell responses [7].

## Materials and Methods

A total of 154 subjects were involved in the study, 68 males and 86 females. The age of patients varied between 25-70 years. All of them were diagnosed as COVID-19 positive by PCR testing. Blood was drawn and plasma was prepared 6-8 weeks after the initial positive testing. None of the patients were on oxygen supply or artificial ventilation.

Blood was centrifuged for 10 000 g for 15 minutes and supernatant incubated at 56^0^C for 30 minutes and centrifuged again. Obtained plasma was diluted 1:100 in PBS for Western blotting experiments.

The mixture of the following proteins was prepared: 1-Recombinant SARS Nucleocapsid protein (Bioss Antibodies Cat.N bs-49002p); 2 - Recombinant SARS-CoV-2 Spike S1 region (Bioss Antibodies, Cat.No. bs-46004P); 3 - Recombinant SARS-CoV-2 Spike RBD (Bioss Antibodies, Cat. No. bs-46003P); 4 - Recombinant SARS-CoV-2 S2 subunit (Ray Biotech, Cat. No. 230-30163). 150 nanogram of each of these proteins were loaded on single line comprised of the gel. Proteins migrate as a bands of the following molecular weights: S1-115-120 kDa; S2-80 kDa, NCP-45-47 kDa and RBD ∼40 kDa

The sodium dodecylsulfate (SDS) gel electrophoresis and Western immunoblotting were carried out as described in our previous studies [8]. After transfer the nitrocellulose membranes were incubated in 3% fat-free milk, then in patient’s plasma (dilution 1:100), washed three times in PBS-Tween, incubated with peroxidase labelled monoclonal anti-human IgG (Abcam, ab99759) and exposed to X-ray films with intensifying screen.

### Statistical analysis

The association between gender and the presence of a protein in the blood was assessed with a two-tailed Fisher exact test. The p-values were corrected for multiple testing with the Benjamini-Hochberg method.

## Results and discussion

The representative images of Western immunoblots are shown on Fig.1. There was a great variability in the antibody response amongst the patients studied. Some of them did not reveal any antibodies against tested proteins, while others demonstrated immune response to only NCP or/and S protein fragments (see Table 1).

**Fig.1.**
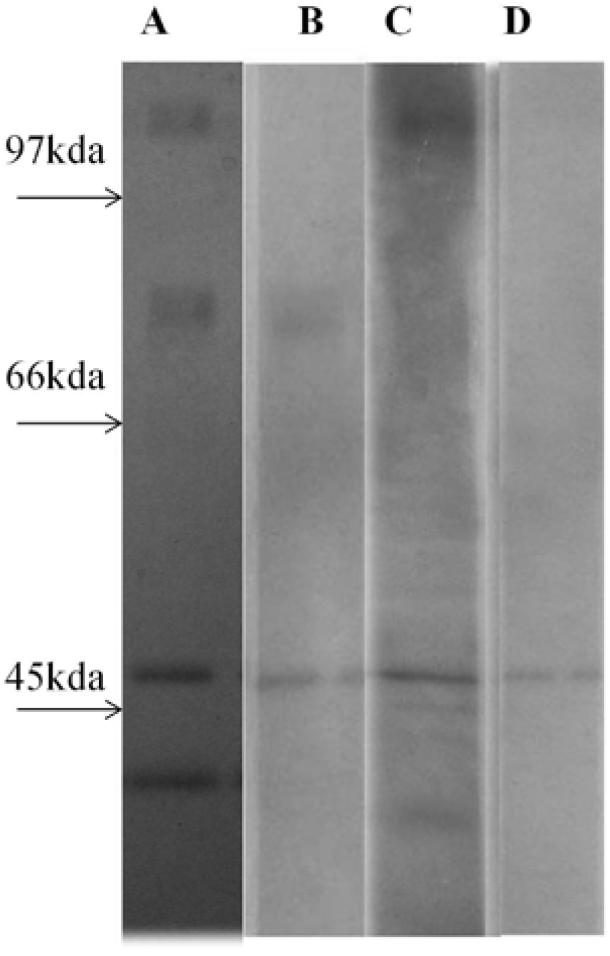
Representative images of: A- Coomassie blue stained gel showing the separation of loaded antigens by ascending order: 1-RBD 40 KDa; 2. NCP 45-47 kDa; 3. S2 fragment 80 Kda and 4. S1 fragment 115-120 kDa. B-D - immunoblots of Patient’s plasma which are characterized with different type of antibodies (lane-B antibodies against S2 and NCP, lane-C antibodies against S1, NCP and RBD and lane-D antibodies only against NCP)

**Table-1.** Distribution of antibody reactivity by antigens in patients. * some patients are producing antibodies to more than one antigen and hence the sum of all positives will exceed the number of patients studied

| Number of patients |  | Absence of antibodies |  | NCP positive |  | RBD positive |  | S1 positive |  | S2 positive |  |
| --- | --- | --- | --- | --- | --- | --- | --- | --- | --- | --- | --- |
| 154 |  | 21 |  | 121 |  | 35 |  | 45 |  | 29 |  |
| Patient number by Gender |  | Absence of antibodies by Gender |  | NCP positive by Gender |  | RBD positive by Gender |  | S1 positive by Gender |  | S2 positive by Gender |  |
| M | F | M | F | M | F | M | F | M | F | M | F |
| 68 | 86 | 13 | 8 | 48 | 73 | 10 | 25 | 12 | 33 | 10 | 19 |

**Fig.2.**
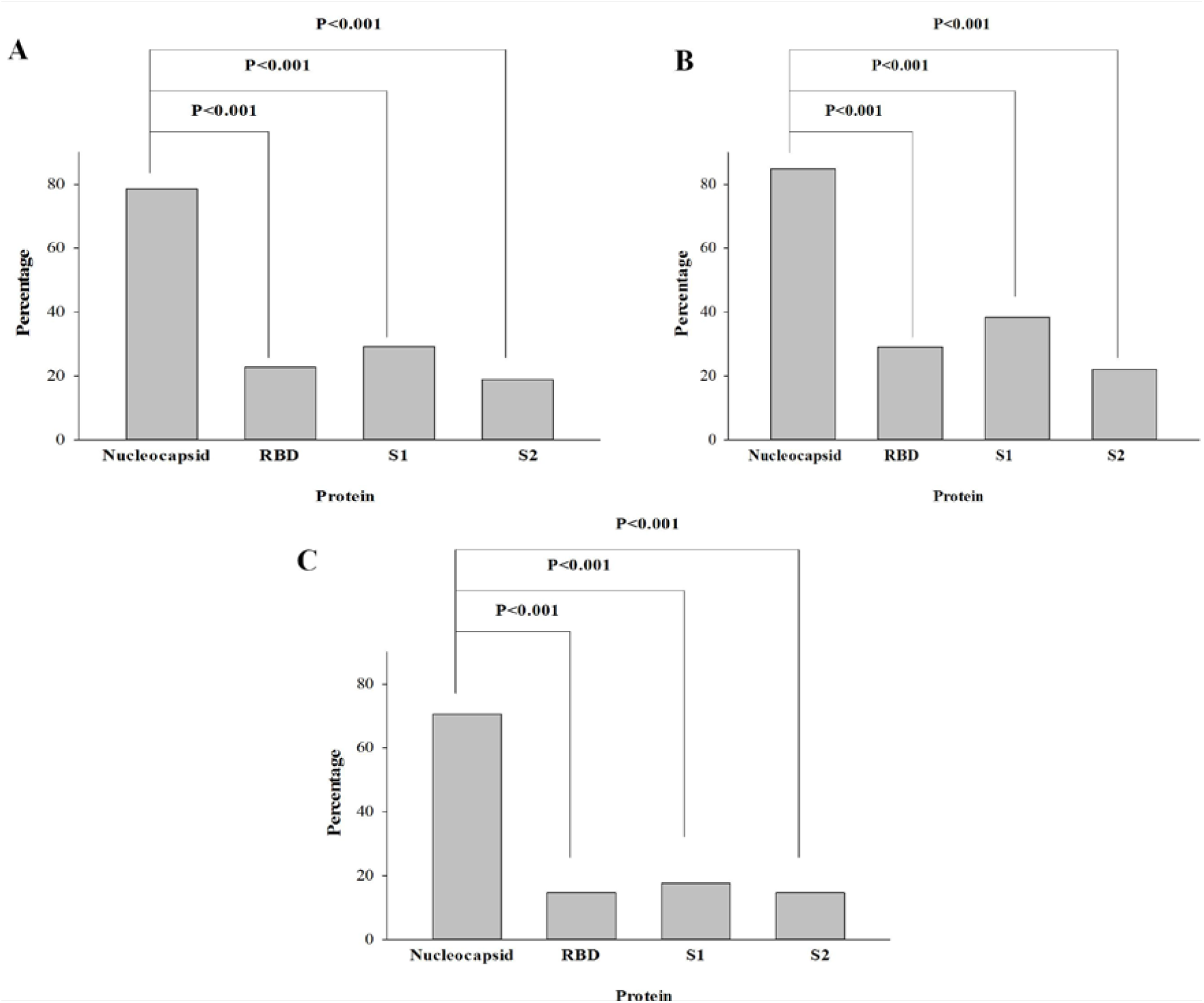
Percentage of patients producing antibodies against to NCP or various fragments of S protein. A-All patients; B- females and C-males. For all cases the antibodies against NCP antibodies are produced in more patients as compared to other antigens studied (adjusted p-value <0.001)

We have compared a gender-specific signature of antibody response to SARS-CoV-2 infection. The percentage of females expressing antibodies to studied antigens in general were higher as compared to males. The presence of the antibodies against S1 fragment is highly associated with gender (corrected P value=0.028), whereas the gender-specific differences for NCP and RBD are significant on one-tailed test only (for both comparisons corrected one-tailed P value=0.035), Fig.3. There are no differences by gender in response to S2 fragment of S protein (P=0.3)

**Fig.3.**
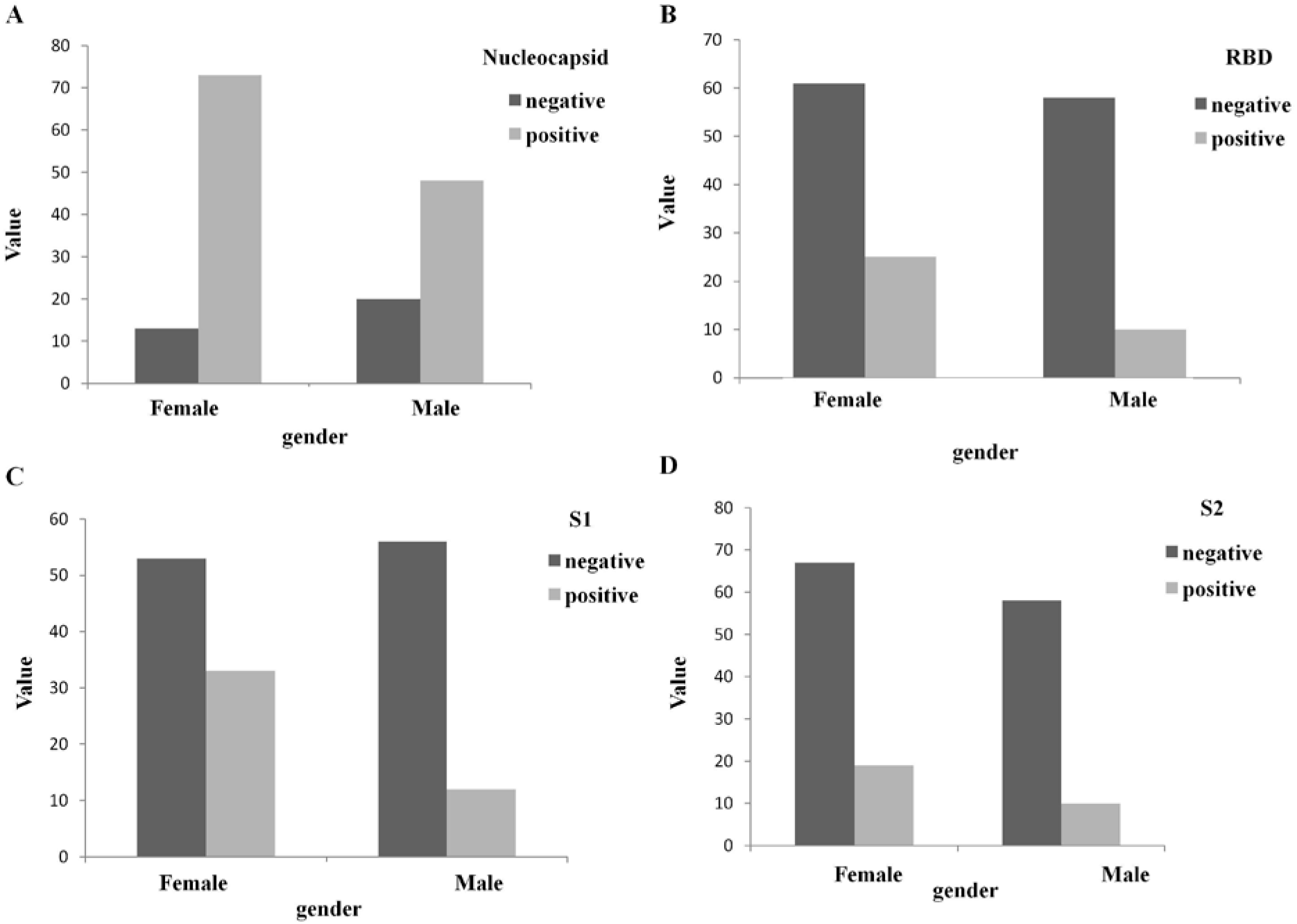
The antibody response to NCP and S protein fragments by gender. A. NCP; B-RBD; C-S1 and D-S2. The significant difference is observed in the case of S1 fragment – higher percentage of females are characterized by the existence of antibodies.

Our obtained data indicate: 1. There is a difference in antibody response to viral proteins; significant majority of patients are producing antibodies against NCP; 2. there is a gender difference in antibody response to SARS-CoV-2 at least in patients with a mild conditions. More females as compared to males are producing antibodies against S1 fragment of S protein. This part of S protein contains RBD – essential region for receptor recognition and virus cell entry and thus antibodies recognizing it would have neutralizing capacities. The distribution of antibodies specifically recognizing only RBD follows the same pattern.

It is well known that across species, females tend to develop a stronger innate and adaptive immune response to infections [reviewed in 9]. In male and female mice with SARS, male mice had a 90% mortality rate, while female mice had a mortality rate of 20% and this sex bias is statistically significant [9,10]. It is supposed that stronger immunity in females increases the reproductive fitness of a species, as mothers are more likely to survive and care for their offspring [9,11]. In support of this suggestion, in animal species where father is responsible for delivering and supporting offspring, upregulation of immunity is observed in males [11].

In humans females have stronger immune response against viral infections than men [12]. Women possess 2 copies of X chromosomes (maternal and paternal), which leads the silencing of one copy of genes in order to ensure an appropriate gene dosage. X chromosome inactivation is cell specific and some cells express the maternal chromosomal copy whereas others express the paternal copy. In female lymphocytes approximately 15 % of X-chromosome genes escape inactivation, leading to biallelic expression with a double dosage (9, 13, 14). Biallelic expression has been shown for CXCR3, TLR7, and CD40L (12, 14). In turn, females possess a diversity of possible immune responses, which provides women with a wider variety of tools with which to fight pathogens [13].

We speculate that more strong antibody response in females could account for the significantly less fatality in spite of higher infectivity of females. Demonstration of such dimorphism in the case of SARS-COV-2 could give basis for the development of selective and efficient therapy separately for mans and women against this viral infection.

## Data Availability

The data which comprise the basic of the manuscript are Western blot images. We are providing only representative images and therefore the images of all original blots will not be available

